# A Partition-Based Group Testing Algorithm for Estimating the Number of Infected Individuals

**DOI:** 10.1101/2021.07.27.21260924

**Authors:** Richard Beigel, Max J. Webber

## Abstract

The dangers of COVID-19 remain ever-present worldwide. The asymptomatic nature of COVID-19 obfuscates the signs policy makers look for when deciding to reopen public areas or further quarantine. In much of the world, testing resources are often scarce, creating a need for testing potentially infected individuals that prioritizes efficiency. This report presents an advancement to Beigel and Kasif’s Approximate Counting Algorithm (ACA). ACA estimates the infection rate with a number of tests that is logarithmic in the population size. Our newer version of the algorithm provides an extra level of efficiency: each subject is tested exactly once. A simulation of the algorithm, created for and presented as part of this paper, can be used to find a linear regression of the results with R^2^ > 0.999. This allows stakeholders and members of the biomedical community to estimate infection rates for varying population sizes and ranges of infection rates.

## Introduction

The COVID-19 pandemic seems to abate in the United States, thanks to the availability of vaccines and testing materials. To date, over 500 million individual coronavirus tests have been performed in the US. Meanwhile, Mexico and Bangladesh, both heavily impacted by the pandemic, have yet to exceed even 8 million tests to date. [1] There continues to be a high demand for testing materials worldwide as production is stretched thin. With the emergence of a more infectious delta variant of the coronavirus [2] there is an impetus to find the number of infected individuals within a given population.

Group disease testing has been used to find the set of positives in a testing population while reducing the tests used since the method was canonized by Dorfman in World War II: testing prospective draftees for syphilis antigens. If the samples from a group of individuals are pooled, and a group tests negative, then those individuals are all negative. In cases where disease rates are low, most of the negatives can be identified easily with few tests. Group testing is therefore an economical alternative to individual testing in these cases. [3,4,5,6]

Scientists in the United States, Israel and Germany have been using group testing since 2020 to detect the presence of COVID-19 in test groups. Computer-ran algorithms have helped to mitigate the challenges to identification presented by multiple infected individuals. [7] One goal of such an algorithm is to count approximately the number of infected individuals using probabilistic sampling. [8,9,10].

In 2020, Beigel and Kasif presented a first version of the Approximate Counting Algorithm (**ACA**): a probabilistic group testing algorithm. This methodology is used to estimate the number of infections by selecting randomly and testing groups of pooled blood samples of varying sizes. [11]

We present a new version of the ACA algorithm which tests each subject exactly once. Then, we simulate both versions of the algorithm.

### Theory

We will show how ⌈log_2_(n+1)⌉ number of tests can be used to approximate the number of infected individuals.

Consider a set S of n test subjects, labeled 1 through n. A small number, k, of those are infected. The subjects are randomly assigned to sets or groups in one of two ways:

- **ACAI:** Sample, randomly (as with a Fisher-Yates “out of a hat” shuffle) with replacement, independent subsets of size ⌈n/2⌉, ⌈n/4⌉, ⌈n/8⌉, ⌈n/16⌉, …, 1.
- **ACAP:** Randomly partition into subsets of size ⌈n/2⌉, ⌈⌊n/2⌋/2⌉, ⌈⌊n/4⌋/2⌉, ⌈⌊n/8⌋/2⌉, ⌈⌊n/16⌋/2⌉, …, 1.

In both cases, test all groups, and count the number of groups where one or more members test positive. The random variable Y denotes the number of positive groups. For ACAI, it has been proved that the Expected Value of 2^Y^ is θ(k). [11] However, the lack of independence makes ACAP hard to analyze mathematically. Instead we verify that relation experimentally and use linear regression to obtain an estimator for k.

Like ACAI, ACAP performs ⌈log_2_(n+1)⌉ tests in parallel.

### Simulation Composition

The simulation we used to test the efficacy of the algorithm was written in Perl, and consisted of 4 different files:

- **GenerateTestData.pl** generates a .csv file with rows of binary strings. Each row is n bits long, and k of those bits are 1s (representing infected individuals). The remaining zeros represent the uninfected. The positions of the 1s and 0s are randomly determined.
- **ACAI.pl** runs a trial of ACAI for each row of the csv file, returning the sample mean of Y and 2^Y^.
- **ACAP.pl** runs a trial of ACAP for each row of the csv file, returning the sample mean of Y and 2^Y^.
- **driver.pl** is responsible for running generateTestData.pl and either ACAI.pl or ACAP.pl for a range of k values. Its output is a .csv file containing a record of the sample mean and sample variance of Y and 2^Y^ for every k.

Using Excel we will graph the recorded k values versus the sample mean(2^Y^) for each algorithm, compare the accuracy of both ACA variants, and determine how closely E(2^Y^) and k are correlated using a linear regression.

The parameters used are n = 1023, and trials per k = 10000. The range of k values used are 1 to 50 (about 0% to 5% infection rate).

## Results

### For k = 1 to k = 50 (about 0% to 5% infected)

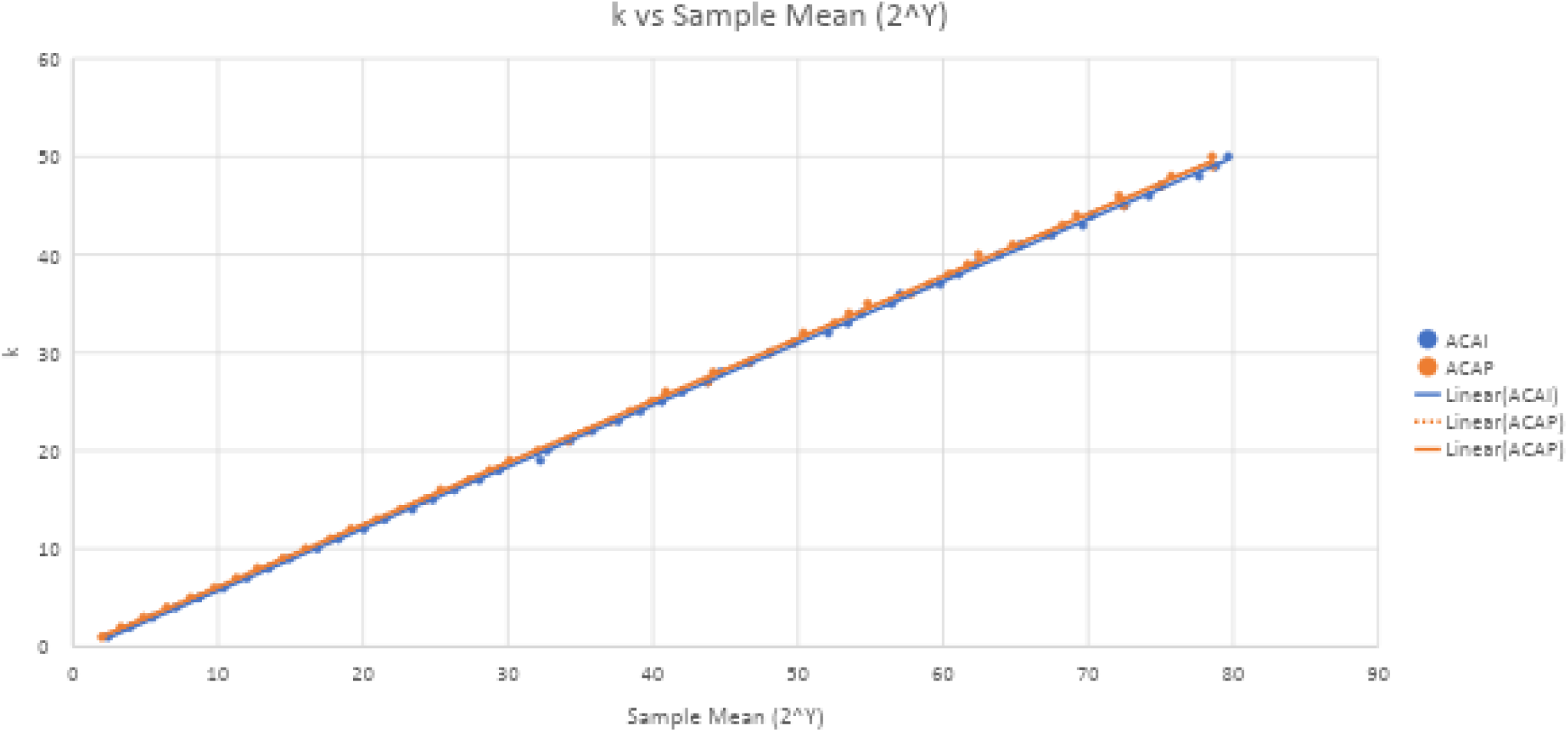

As demonstrated in the figure above, there is a strong correlation between k and the Sample Mean(2^Y^). There are no outliers. The linear regressions of both algorithms have acceptable R-values of .9998 and .9996, respectively. This suggests either of these linear equations would be useful estimators for k at low (<5%) infection rates.

### For k=1 to k =100 (about 0% to 10% infected)

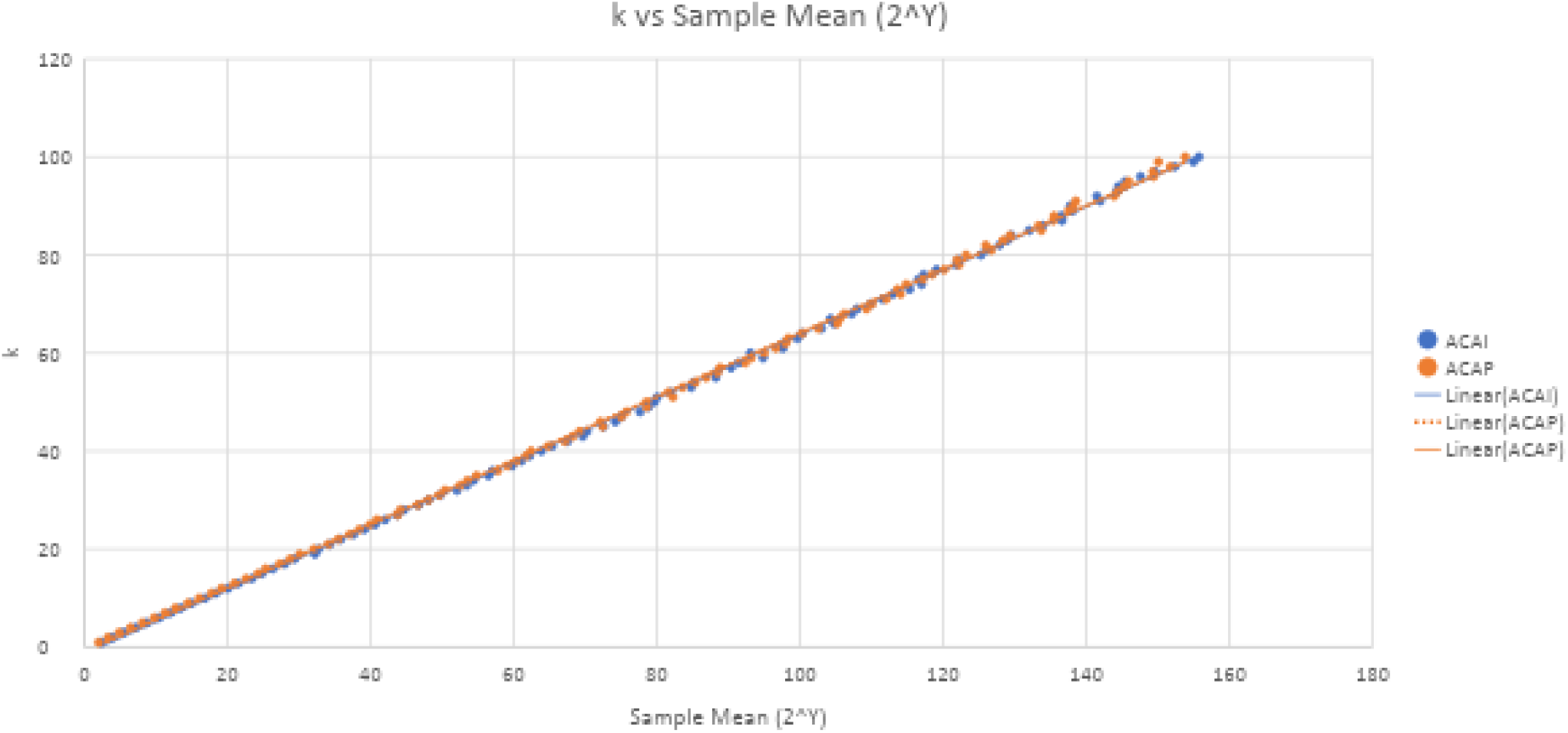

When we widen the range of values, the R-squared values decrease somewhat, with both linear regressions looking equally effective with an R-squared value of .9995. The trendline is tightest bound around the middle of the graph. Despite this, the correlation is still strong throughout. These formulae would be acceptable estimators for k, but it’s worth noting how the linear formulae differ between the 1 to 50 and 1 to 100 ranges for k. Both the multiplicative and additive constants are higher.

## Discussion

The COVID-19 pandemic remains ever-present in humanity’s concern worldwide. Approximating the number of infected individuals can inform decision-makers responsible for reducing the spread of COVID-19. For example, an approximate count can be the tipping point for whether schools reopen in-person or live events are held. This form of batch testing can be both economical and revelatory. While EY in this case was a useful estimator for log_2_(k), E(2^Y^) is biased. Exponentiating the relationship between EY and log_2_(k) will not yield directly useful results. It was the linear regression of k versus 2^Y^ that proved to be the best estimator for k. Detecting the presence of the disease as early as possible coincides with the goal to approximately count the infected at low prevalence.

The code used for this simulation can be freely accessed and downloaded at https://github.com/maxjwebber/covidcounting. Users should install Perl, move the files to the same directory and run driver.pl with the following command line parameters. Usage is:

driver.pl -n [number of subjects] (required) -kmin [smallest number of infected] (optional) -kmax [largest number of infected] (optional) -trialsperk [number of trials to average for each number of infected] (optional).

For example, driver.pl -n 1023 -kmin 1 -kmax 50 -trialsperk 10000

We recommend a lowest k value of 0 or 1 for the best fit (the program will warn a user who uses a different value). A pair of csv files, ACAP_results.csv and ACAI_results.csv will be created with the results of the individual k values, and the linear regression formulae will be written to linear_regression.csv along with the R-squared values for each. Use the formula that corresponds to the algorithm you use in the lab.

## Data Availability

Data used for graphs is available freely in Github repository with simulation code.

https://github.com/maxjwebber/covidcounting

